# Epidemiology of Venous Thromboembolism in SARS-CoV-2 Infected Patients: A Systematic Review and Meta-Analysis

**DOI:** 10.1101/2020.08.28.20184028

**Authors:** Souvik Maitra, Dalim K Baidya, Sulagna Bhattacharjee, Rahul K Anand, Bikash R Ray

**Author notes:** Correspondence to: Dr. Sulagna Bhattacharjee MD, DNB, DM, Assistant Professor, Room No: 5011, Teaching Block, Department of Anaesthesiology, Pain Medicine & Critical Care, All India Institute of Medical Sciences, Ansari Nagar, New Delhi-110029 Email id.

## Abstract

Early reports from China and Europe indicated that incidence of venous thromboembolism in COVID-19 patients may be high. In this meta-analysis of observational studies was designed to know worldwide prevalence of thromboembolic events in COVID-19 patients. Primary outcome of our review was to assess the proportion of patients with VTE. Secondary outcomes were to assess the proportion of patients’ with DVT and proportion of patients with PE. Random effect meta-analysis model with restricted maximum likelihood estimator was used for all analysis. Pooled proportion with 95% confidence interval (95% CI) and heterogeneity (I^2^) was reported for all outcomes. Data of 5426 patients from n=19 articles were included in this systematic review and meta-analysis. Incidence of VTE (95% CI), PE (95% CI) and DVT (95% CI) was 23 (10-36) %, 12 (6-17) % and 15 (8-23) %. We have found a high but incidence of thromboembolic events in COVID-19 patients. Further well-designed studies are required in this area to identify true incidence and risk factors of it.

Key Messages
- This meta-analysis of observational studies was designed to know worldwide prevalence of thromboembolic events in COVID-19 patients.
- Data of more than 5000 patients from 19 observational studies were analyzed in this meta-analysis.
- Incidence of venous thromboembolism may be as high as 36% in these patients.

## Introduction

Since the diagnosis of first confirmed case of SARS-CoV-2 infection in December 2019, more than 15,000,000 people have been affected and more than 600,000 people have died from it [1]. SARS-CoV-2 infection is associated with a hypercoagulable state possibly due to endothelial dysfunction secondary to the infection [2]. Early reports from China and Europe indicated that incidence of venous thromboembolism in these patients may be high [3, 4]; however, whether there is a difference between COVID-19 and non-COVID-19 patients is controversial [5]. However, most of the individual studies reporting incidence of thrombotic event were of small sample size and interpretation of individual study results difficult. Moreover, there is wide variability in the reported incidence of thromboembolic event in the individual studies. So, this meta-analysis of observational studies was designed to know worldwide prevalence of thromboembolic events in COVID-19 patients.

## Methodology

This systematic review and meta-analysis were conducted as per PRISMA methodology [6]. All peer-reviewed published studies, including RCTs, prospective cohort studies and retrospective studies, where incidence of lower limb venous thromboembolism was reported in SARS-CoV-2 infected adult patients were included in this systemic review and meta-analysis. Non-peer reviewed article, case reports, case series of less than 10 sample size and studies conducted in pediatric patients were excluded from this review.

Eligible articles for this systematic review and meta-analysis were searched from PubMed and PubMed central with the key words “venous thromboembolism and SARS-CoV-2”, “venous thromboembolism and COVID”, “venous thromboembolism and COVID-19”, “venous thromboembolism and coronavirus”, “VTE and COVID-19”, “VTE and SARS-CoV-2”, “VTE and coronavirus”, “VTE and COVID”, “pulmonary embolism and SARS-CoV-2”, “pulmonary embolism and COVID”, “pulmonary embolism and COVID-19”, “pulmonary embolism and coronavirus”, “PE and SARS-CoV-2”, “PE and COVID”, “PE and COVID-19”, “PE and coronavirus”, “DVT and SARS-CoV-2”, “DVT and COVID”, “DVT and COVID-19”, “DVT and coronavirus”, “deep venous thrombosis and SARS-CoV-2”, “deep venous thrombosis and COVID”, “deep venous thrombosis and COVID-19”, “deep venous thrombosis and coronavirus”, from inception to 26^th^ July 2020.

Required data from the eligible studies were extracted by two independent authors from the and then all data were tabulated in a predefined Microsoft Excel™ (Microsoft Corp., Redmond, WA) data sheet. All data were cross-checked by the third review author (SM). Risk of bias in individual studies were assessed by two independent authors (SM & SB) as per ‘Risk of Bias in Non-randomized Studies of Interventions’ methodology.

Primary outcome of our review was to assess the proportion of patients with VTE. Secondary outcomes were to assess the proportion of patients’ with DVT and proportion of patients with PE. Random effect meta-analysis model with restricted maximum likelihood estimator was used for all analysis. Pooled proportion with 95% confidence interval (95% CI) and heterogeneity (I^2^) was reported for all outcomes. All analyses were performed with *metafor* package in R (R version 3.6.1, R Development Core Team, 2010; R Foundation for Statistical Computing, Vienna, Austria) in *Jamovi* platform. Association between outcome of interests (incidence of VTE/ PE) and demographic and clinical characteristics of the patients were planned by mixed effect linear meta-regression model if more than 10 studies were available for analysis. Standardize regression coefficient with standard error (SE) and unadjusted R^2^ (amount of the heterogeneity accounted for that particular model) for each model were also estimated.

## Results

Initial database searching revealed 2259 articles and another 12 articles were retrieved from other sources. After duplicate removal 287 articles were screened from title and abstract. Finally, data of 5426 patients from n=19 articles were included in this systematic review and meta-analysis [2-5, 7-21]. Two studies were conducted in China [4, 16], three were in United States [13, 20, 21], and rest were conducted in Europe [2, 3, 5, 7-12, 14, 15, 17-19]. Only five studies were prospective in design [2, 3, 5, 10, 15] and rest 13 were retrospective. Routine DVT screening was performed only in 3 studies [15, 19, 20] and screening on the basis of clinical symptoms were incorporated in rest of the studies. Thromboprophylaxis protocol was described in 13 studies, no thromboprophylaxis was used in one study [4] and 4 studies [2, 5, 12, 13] didn’t describe thromboprophylaxis protocol. All included studies were high risk of biases because of variable thromboprophylaxis protocol, screening protocol, follow-up duration and obvious selection bias. Details of the included studies reported in table 1.

Incidence of VTE was reported in 10 studies and pooled estimate (95% CI) was 23 (10-36) %; however significant heterogeneity (I^2^=98.8%, p<0.001) and publication bias (p=0.001, regression test for funnel plot asymmetry) were found. Fourteen studies reported incidence of PE and pooled estimate (95% CI) was 12 (6-17) % and significant heterogeneity (I^2^=99.3%, p<0.001) and publication bias (p<0.001, regression test for funnel plot asymmetry) was reported. Pooled estimate (95% CI) of DVT was 15 (8-23) % and it was reported in 16 studies. Significant heterogeneity (I^2^=98.8%, p<0.001) and publication bias (p<0.0001, regression test for funnel plot asymmetry) were also found in this outcome also. Neither mean age of the recruited patients nor proportion of patients mechanically ventilated at baseline was significant moderator of any of the above-mentioned clinical outcomes. Male sex was significantly associated with development of PE [effect estimate (SE) 0.007 (0.003), R^2^= 29.6%, p=0.02].

## Discussion

In this meta-analysis we have found an overall high incidence of VTE, PE and DVT in COVID-19 patients across various disease severity and male sex was possibly associated with development of PE in these patients. Previous reports suggested that VTE related events in acutely ill medical patients is around 2 per 100 patients even with thromboprophylaxis [22]. Another multicentric study reported that 7.7% critically ill medical-surgical patients developed venous thromboembolism despite pharmacological thromboprophylaxis [23].

True incidence of thromboembolic events in COVID-19 patients is expected to be even higher as routine DVT/ PE screening at serial interval was not used in most of the studies. Secondly, significant numbers of patients were still in the hospital at the time publication of the study. Various thromboprophylaxis regimen was used in different studies which could have also impacted incidence of thromboembolic event. We have found significant amount of heterogeneity in all pooled thromboembolic events which could be due to ethnic variation of patients, variable screening protocol and thromboprophylaxis protocol, disease severity and patients’ comorbid illness etc.

To conclude, we have found a high but incidence of thromboembolic events in COVID-19 patients. Further well-designed studies are required in this area to identify true incidence and risk factors of it.

## Data Availability

Data will be available from the corresponding author on reasonable requests.

---

**Figure 1:**
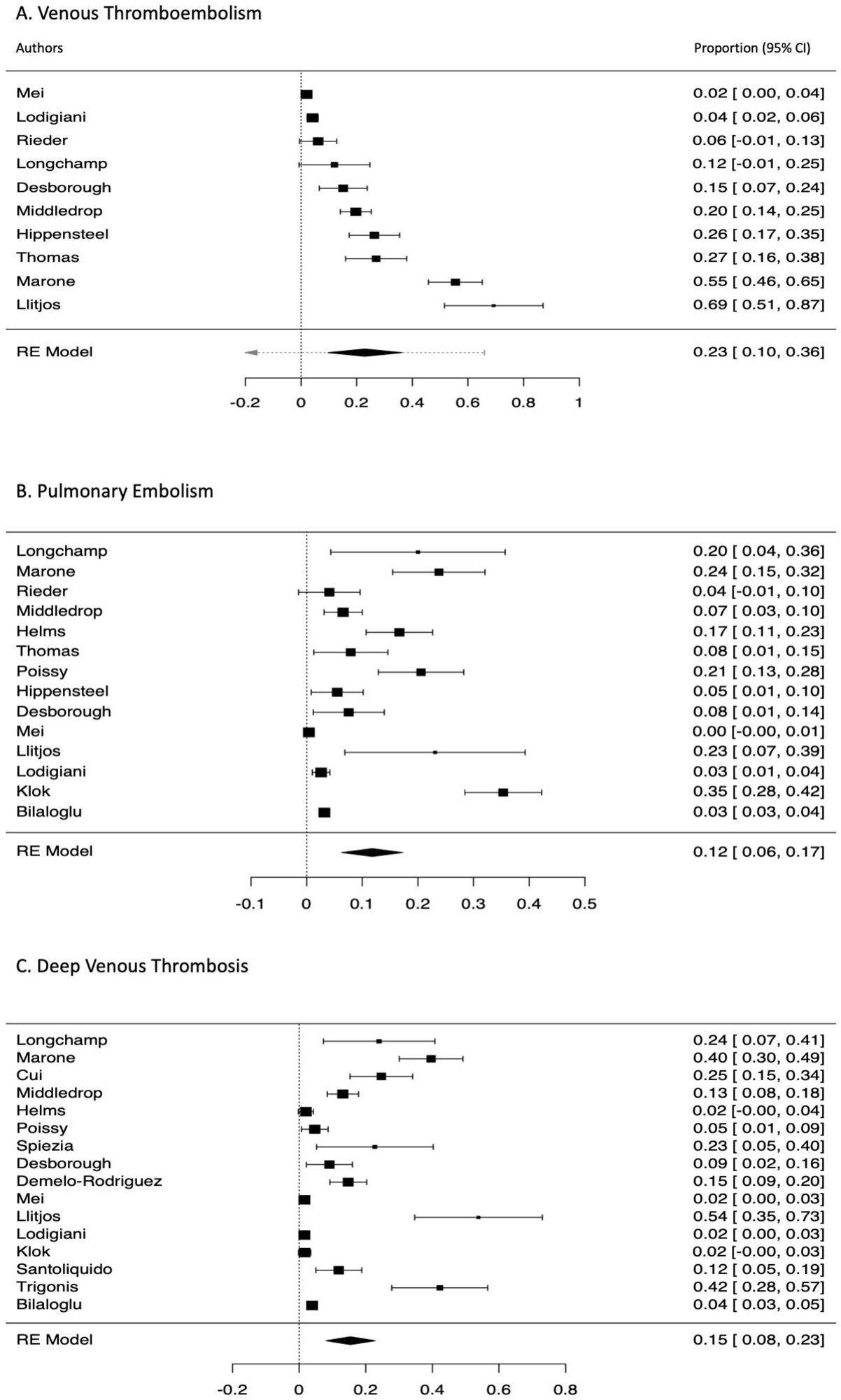
Forest plot showing pooled estimate of venous thromboembolism (Upper), pulmonary embolism (Middle) and deep venous thrombosis (Lower)

